# Clinical audit on laboratory diagnosis of malaria parasites at Jimma University Medical Center laboratory, Southwest Ethiopia

**DOI:** 10.1101/2025.02.17.25322418

**Authors:** Dechasa Girma, Adane Hailu, Chalachew Abebe, Hawa Said, Hamda Mohamoud, Sintayehu Asaye

## Abstract

1.

**Background:** Microscopic diagnosis using stained blood films is usually regarded as the gold standard and most acceptable method of diagnosis. There is no clinical audit performed at the Clinical Laboratory of Jimma University Medical Center, which aims to audit malaria diagnosis; therefore, this audit assesses the pre-analytic, analytic, and post-analytic processes that can affect the accuracy and quality of malaria diagnosis in this laboratory against the established acceptable standards of malaria diagnosis.

**Method:** We conducted a clinical audit at the Jimma University Medical Center Laboratory from January 2-20, 2025, using direct observation of the procedure that laboratory staff follow in blood collection, fixation, blood smear preparation, microscopic examination, reporting, and documentation of malaria diagnosis results, and also by re-examining the diagnosed slides to assess the skill of laboratory staff by maintaining confidentiality of the data.

**Results:** A total of 26 quality standards were used. It is known that 56.3 percent of the samples are collected from capillaries and 43.7 from veins. Approximately 78 percent of the samples were collected from properly rubbed fingers and 22% from fingers that were not rubbed adequately. All the blood samples collected with an anticoagulant were mixed properly before smearing (n=35), 78.8 % of the slides were preliminarily visualized using a high-power (100x) objective of the microscope, and only 17.5 % of the slides were visualized using a 10x objective.

**Conclusion:** Proper Disinfection, mixing the anti-coagulated blood well, and fixing thin blood films for the recommended length of time were areas of good practice. On the other hand, failure to report parasitic load and proper and sequential use of types of objectives to examine blood films and prepare smears of good quality were some of the highly violated standards that need consideration.

## 2. Introduction

Malaria is a parasitic disease transmitted by vectors and caused by protozoan parasites from the *Plasmodium* genus, primarily affecting tropical and subtropical regions worldwide(1). *Plasmodium* comprises over 200 species that infect mammals, birds, and reptiles(2). The five recognized species of *Plasmodium* responsible for malaria are *Plasmodium falciparum, Plasmodium vivax, Plasmodium malariae, Plasmodium ovale*, and *Plasmodium knowlesi(3)*.Of the genus *Plasmodium* that causes malaria disease in humans, *plasmodium falciparum* causes severe life threatening infections (4).

Globally, it is estimated that 3.2 billion people are at risk of contracting malaria annually(5). Although malaria is both preventable and treatable, it continues to have a significant impact on global health, especially among pregnant women and children in both rural and urban regions(6).

A WHO report indicated that the majority of the world’s malaria burden is concentrated in sub-Saharan Africa, which also records the highest number of cases and fatalities, and this ongoing health crisis is not only harming people’s well-being, but also hindering the economic progress of numerous developing nations, especially in sub-Saharan Africa(7).

About 90% of the world’s malaria deaths are estimated to occur in sub-Saharan Africa, where the majority of the infections are caused by the species Plasmodium falciparum, which was known to cause severe manifestations (8). Although most of severe cases of malaria are caused mainly by Plasmodium falciparum (P.falciparum), but can also be caused sometimes by Plasmodium vivax and mixed infections(9).

Malaria is a significant health issue in Ethiopia, being a leading cause of illness and death for many years. Data from the Ethiopian Federal Ministry of Health shows that 75% of the country is affected by malaria, with approximately 68% of the population residing in areas vulnerable to the disease(10), and it is still a major public health concern, and one of the leading causes of morbidity and mortality among adult populations(3). Epidemics typically occur in the highland or highland border regions, particularly in areas located 1,000 to 2,000 meters above sea level(10).

The Southern Nations, Nationalities, and Peoples’ Region (SNNPR) had the highest malaria prevalence in Ethiopia at 16.17%, followed by Oromia Regional State at 13.11% and Amhara Regional State at 12.41%(11). A hospital-based study in Ethiopia found that 7.7% of blood film examinations conducted in that laboratory facility were positive for malaria, and among the confirmed cases, 47.2% were due to *P. vivax*, 45.6% were caused by *P. falciparum*, and 7.2% were mixed infections(12), which is comparable to the result of a study performed on the other area of the country (5%) in terms of general prevalence, but varying in distribution of the species, with p. falciparum accounting for 54% of the cases and p.vivax accounting for 44% of the cases with no finding of mixed cases(13). This shows the variability in the distribution of species of malaria parasites among different areas of the country.

Jimma is an area in the Oromia region of Ethiopia that is known to be endemic for malaria(9). The prevalence of urban malaria in Jimma Town was 5.2% according to a study done previously, with a higher malaria prevalence rate among under-five children (11%)(14). Another study conducted at Jimma University Medical Center(JUMC) from May 2013 to 2016 indicated that the prevalence of confirmed severe and complicated malaria was 2.6 % (144), and among the confirmed cases of severe and complicated malaria, the case mortality rate was 0.6% (32 deaths), while the case fatality rate stood at 22.2% (32 out of 144)(9).

The clinical laboratory plays a vital role in the initial diagnosis, tracking the effectiveness of anti-malarial treatments, detecting complications from severe malaria, and identifying the development of drug resistance(15). The World Health Organization (WHO) recommends parasitological diagnosis for all patients suspected of having malaria, as relying solely on clinical diagnosis can be inaccurate and increase the risk of antimalarial drug resistance(16), and of the mechanisms of malaria diagnosis, microscopic diagnosis using stained blood films is usually regarded as a gold standard and the most acceptable way of diagnosis. Most clinical laboratories diagnose malaria by stained blood films using Giemsa’s stain more frequently and Wright’s stain, rarely(15).

This audit aimed to assess the pre-analytic, analytic, and post-analytic processes that can affect the accuracy and quality of the malaria microscopy results. These processes include sample collection methods, blood smear preparation, fixation, staining procedure, documentation, and the skill and culture of the laboratory staff while examining blood films and identifying malaria parasite species in the Clinical Laboratory of JUMC against the established acceptable standards.

## 3. Method

The clinical audit was conducted at the Jimma University Medical Center Laboratory (January 2025 G.C) using direct observation of the procedure that laboratory staff follow in blood collection, fixation, blood smear preparation, microscopic examination, reporting and documentation of malaria diagnosis results, and also by re-examining the diagnosed slides to assess the skill of laboratory staff by keeping the confidentiality of the data. Finally, descriptive analysis was done using SPSS software version 26, the findings are presented in statements and figures.

### Sample size

A total of 80 blood sample collections, preparations, and laboratory investigations over 15 days were audited by postgraduate students of clinical chemistry and hematology departments. Of the 80 blood samples, 45 were collected from capillaries using blood collection lancets and 35 from veins using an anticoagulant tube.

### Standards

The following were the areas of consideration when conducting the audit:

### Collection of blood samples

#### For venous blood

To obtain venous blood, selecting the appropriate vein (the median one) and using the appropriate syringe gauge size according to the size of the vein, then mixing gently with anticoagulant to avoid clotting of blood. Within 30 minutes of collection, placement of an adequate amount of blood on a clean slide, and making a good blood film were used as the main standards.

#### For capillary blood

Massaging the puncture site 5-6 times to increase blood flow and cleansing with 70% alcohol, then making a puncture of 2-3 mm deep with a sterile lancet, removing the first drop of blood, placing a sufficient amount of blood on a clean slide, and making a tongue-shaped blood film that covers 3/4^th^ of the slide and allowing it to air dry were considered.

#### Staining procedure for Giemsa’s stain

The audit on the staining procedure made its target of covering the blood smear completely with Giemsa stain and the time of staining. To perform this first step, preparing a working Giemsa solution from the stock solution is essential.

At the time of this audit, a 10% Giemsa working solution was being used, and he time of staining in this case was about 10 minutes. Then after it is dried well, staining it appropriately with prepared Giemsa by following standard operating procedure (SOP) was considered.

#### Quality of stained blood smears prepared

In addition to the distribution of red blood cells during smear preparation, the quality of the stained blood films was assessed following standards. Standards were set for blood films stained with Giemsa staining solution. On microscopic examination of the blood films, the characteristic color of each blood cell was used as a standard. Accordingly, the blue cytoplasm and red-purple chromatin dots of the diagnostic stages of malaria parasites (Trophozoites, Schizonts, and Gametocytes) were considered. Moreover, appearance of red blood cells (pale pink or reddish, with pale center), neutrophils (light pink or purple granules), lymphocytes (dark purple nucleus), eosinophils (bright orange to red-orange granules), basophils (dark purple granules in the cytoplasm), and platelets (pale blue granules) were the major targets for assessing the quality of the blood films.

#### Microscopic examination of the blood film and Objectives of the Microscope used

The audit on the microscopic examination of the stained blood films was conducted following standards that initially assessing the overall distribution and quality of the blood film with 10x objective, identifying the different blood cells, abnormalities, and presence of malaria parasites inside red blood cells using 40x objective and finally, addition of a drop of immersion oil on stained blood film and examination of the preparation by oil immersion(100x) objective for final confirmation, were considered.

#### Reporting Systems

The audit on the reporting system of the malaria diagnosis results was based on the standards that included a minimum of 100 fields of microscopic observation, examining for a minimum of 5 minutes microscopically before reporting the results and reporting negative slides, as “No hemoparasite was seen”. Moreover, reporting the type of malaria parasite and the observed stages of the parasite in the case of *Plasmodium falciparum* together with parasitic load for slides with confirmed malaria positivity, were also considered.

#### Documentation system

The audit on documentation of malaria diagnostic results was based on the standard that the documentation includes Patient Information (Full Name, age, sex, patient ID/record Number), Laboratory Details (Laboratory name and location, lab Technician name), date of lab analysis, clinical Information, and test Results.

## 4. Results

A total of 26 quality standards were used to assess overall malaria diagnosis. It was known that 56.3 percent of the samples were collected from capillary and 43.7% from veins. Approximately 78 % of the samples were collected from properly rubbed fingers, 22% from fingers that were not rubbed adequately, and 98.8% of the capillaries (fingertips) were properly cleaned with 70% alcohol before puncture. All malaria diagnosis results (including the rechecked slides) were recorded and documented according to the standard (n=80).

All of the blood samples collected with an anticoagulant were mixed properly before smearing (n=35) and 88.9% of the blood samples collected from capillaries were collected after wiping the first drop of blood (n=45), and 11.1 % without removing the first drop of blood. All smears were fixed for the length of time recommended by the standard (n=80), and more than half (66.3%) of the thin smears did not have optimal length **(Figure 1)**. It took less than 2 minutes, more than 5 minutes, and between 2 and 5 minutes for 10%, 16.2%, and 73.8% of the professionals, respectively, before reporting as “No hemoparasite seen” **(Figure 2)**.

**Figure 1:**
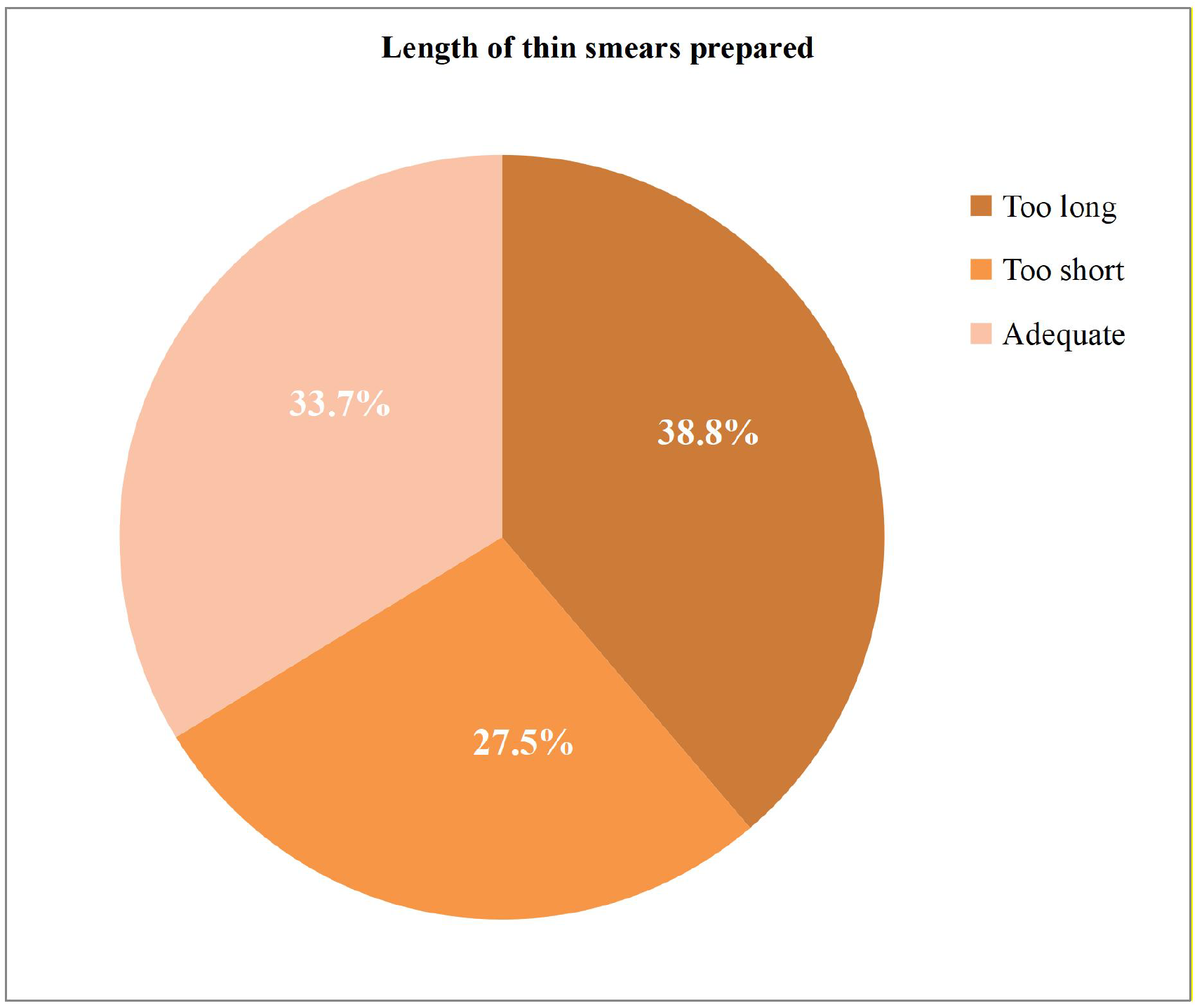
Adequacy of the length of thin smears prepared by staff (n=80).

**Figure 2:**
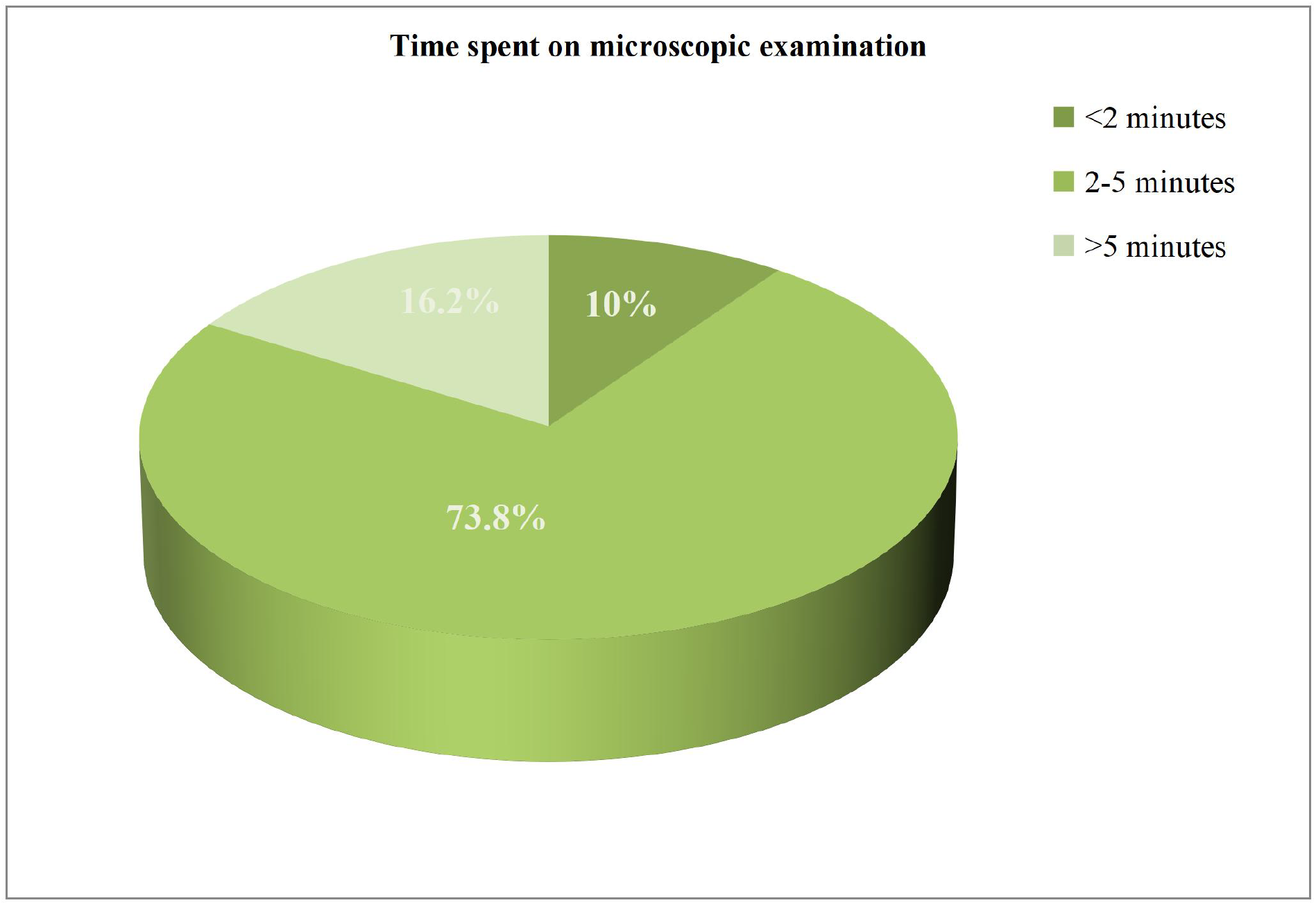
Time spent on microscopic examination by staffs before reporting as “No hemoparasite seen” (n=80).

Moreover, the use of clean or new slides and the placement of a sufficient amount of blood on a slide were 95 % and 80 %, respectively, and all the prepared smears were air-dried. Regarding the staining of blood films, all of the blood samples were covered well with Giemsa stain and 78.8 % of the slides were preliminarily visualized using the high power (100x) objective of the microscope, and only 17.5 % of the slides were visualized by using 10 x objective **(Figure 3)**. Laboratory professionals reported the species of the parasite and its developmental stage on all of the samples that were positive (n=17).

**Figure 3:**
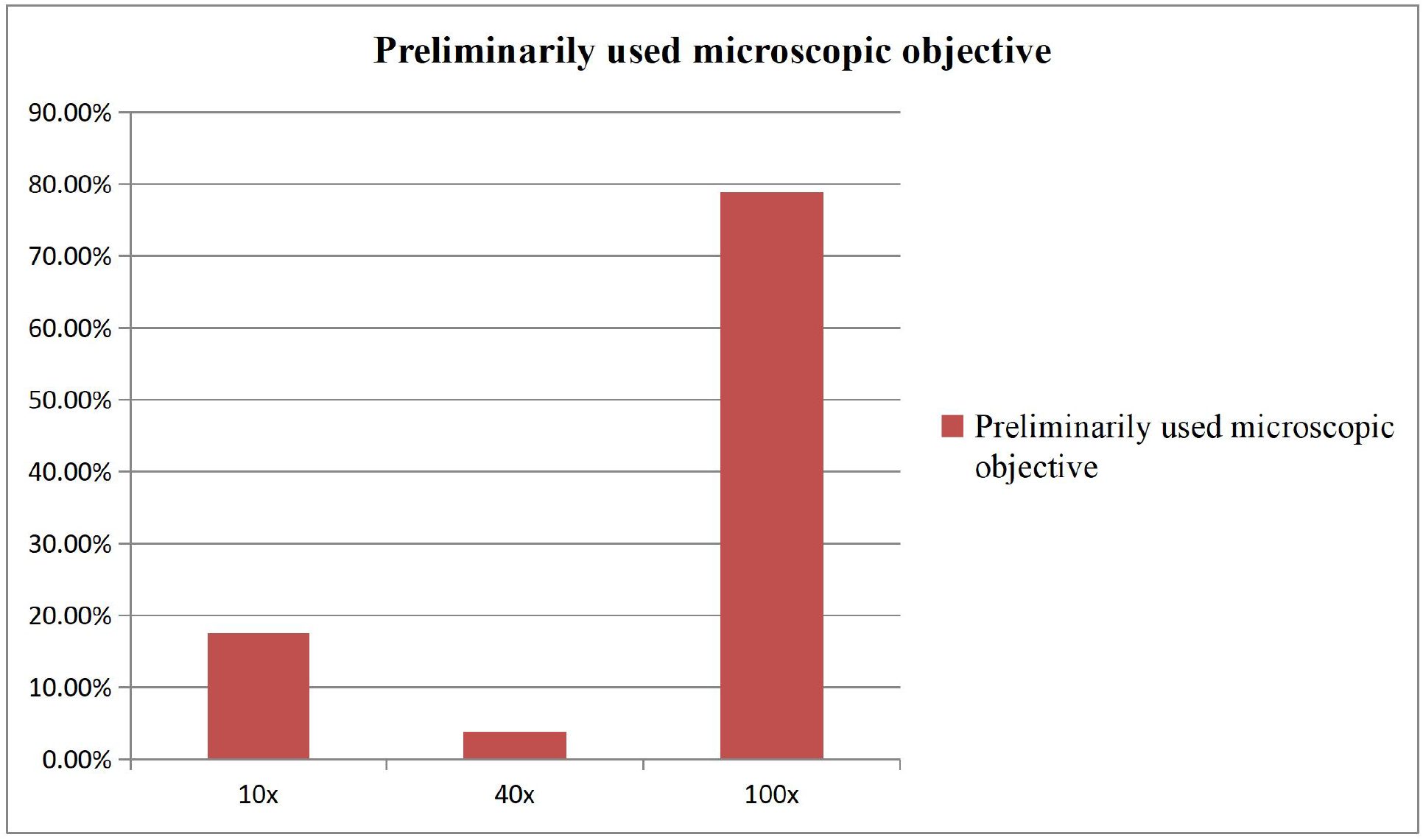
Microscopic objective preliminarily used by staff during blood film examination (n=80).

Moreover, parasitic load is not reported in all of the positive cases. Additionally, re-checked slides (n=62) showed that all positive slides rechecked were true positives, and also there were no false negatives among the re-examined slides.

## 5. Discussion

This audit has revealed the typical features and acceptable approaches to malaria diagnosis in the Jimma University Medical Center Laboratory. In this audit, it was found that fingertips were not rubbed properly in 22.2 % of blood samples collected, but capillary blood collection guideline recommends gently massaging the finger from the tip towards the hand 5-6 times so as to improve blood circulation, but avoiding application of too much pressure is important according to this guideline(17), as excessive squeezing may lead to an accumulation of tissue fluids in the area, which could impact the accuracy of the results.

This audit showed that 11.1 % of the capillary blood samples were collected without removing the first drop of blood, which showed better adherence than the result of a clinical audit conducted at University of Gondar Referral Hospital(38%), but only 33.8% of the thin smears had an adequate and optimal length in this audit, which is much lower than the previous audit(15). Standard blood film preparation guidelines recommend that the smear should cover two-thirds of the base slide length and should have an oval feathered end that allows to create a single layer of cells for optimal view of cellular morphology and diagnosis of hemoparasites(18).

In this audit, both thin and thick smears were prepared on 98.8% of the samples (n=80). Well-made thick blood films are evenly spread, have 10–15 white blood cells (WBCs) per high-power field (HPF) on average, and good quality thin films have a feathered edge with a monolayer of red blood cells (RBCs) that help in identifying malaria species, including trophozoites in mixed infections, estimating parasitemia levels and they must be spread using the unchipped edge of a ground-edge slide (19).

The results of this audit showed that only 3.8 % of the slides were visualized using the high power (40X) objective of the microscope before being examined by the oil immersion objective. However, WHO malaria microscopy guideline advises that blood smears should be analyzed under a microscope using both the 40X and 100X objective lenses(20). Moreover, the microscopic examination time before releasing the slides as negative for hemoparasite, had variability among laboratory technicians followed in this audit.

In this audit, parasitic load was not reported in all the positive cases found. However, WHO recommends that whenever malaria parasite is detected, particularly Plasmodium falciparum or Plasmodium knowlesi, the percentage of parasitized cells should be quantified using a thin film preparation, examining a minimum of 1,000 red blood cells across various areas of the film as well as further quantification on a thick film, as severity of parasitemia can significantly affect the choice of treatment (20).

## 6. Conclusion

Disinfection of the blood collection site, mixing the anti-coagulated blood well before preparing blood film, and fixing thin blood films for the recommended length of time were the areas of good practice identified in this audit. However, failure to report parasitic load and proper and sequential use of the types of objectives to examine blood films and preparing smears of good quality are some of the highly violated standards that need a consideration.

## 7. Recommendation

Appropriate actions such as training the staff to improve their skills and setting rules that every staff should follow to implement an adequate reporting system to provide the actual information regarding the status of the sample owner should be considered.

## Data Availability

Data will be available upon convincing request from the corresponding author.

https://www.cdc.gov/malaria/hcp/clinical-guidance

## Declarations

## Ethical approval and consent to participate

Ethical statement was waived by Jimma University Medical Center Laboratory management, as it was a clinical audit and no human subjects were involved.

## Consent for publication

Not applicable

## Availability of data and materials

Data will be available upon convincing request from the corresponding author.

## Competing interests

The authors declare that they have no competing interests.

## Funding

The authors did not receive any funding to support the conduct of this audit.

## 8. Author contributions

All listed authors provided intellectual contributions and made critical revisions to this audit report; AH and DG have contributed to study design, data analysis and interpretation; CA, HS and HM contributed to audit data acquisition; SA has contributed to the conceptualization and has critically participated in the supervision from the inception to the end of the audit. Besides that, all authors certified the contribution description and approved the final version of the manuscript.

## 9. Acknowledgement

We thank the laboratory managers, quality and safety officers, and technologists at Jimma University Medical Center for their invaluable support during the data collection process.

## Notes

**Financial support and conflict of interest disclosure:** The authors of this clinical audit have no conflict of interest and have received no funding for the audit.

### Competing Interest Statement

The authors have declared no competing interest.

### Author Declarations

Ethical committee of Jimma University Medical Center Laboratory waived ethical approval for this work

## References

1. Escalante AA, Pacheco MA. Malaria molecular epidemiology: an evolutionary genetics perspective. Microbiology spectrum. 2019;7(4):10.1128/microbiolspec.ame-0010-2019. https://doi.org/10.1128/microbiolspec.ame-0010-2019

2. Singh B, Daneshvar C. Human infections and detection of Plasmodium knowlesi. Clinical microbiology reviews. 2013;26(2):165–84. 10.1128/cmr.00079-12

3. Delil RK, Dileba TK, Habtu YA, Gone TF, Leta TJ. Magnitude of malaria and factors among febrile cases in low transmission areas of Hadiya zone, Ethiopia: a facility based cross sectional study. PLoS One. 2016;11(5):e0154277. 10.1371/journal.pone.0154277

4. Antony HA, Parija SC. Antimalarial drug resistance: an overview. Tropical parasitology. 2016;6(1):30–41. DOI: 10.4103/2229-5070.175081

5. Ba EH, Baird JK, Barnwell J, Bell D, Carter J, Dhorda M, et al. Microscopy for the detection, identification and quantification of malaria parasites on stained thick and thin blood films in research settings: procedure: methods manual. 2015. ISBN: 978-92-4-154921-9

6. Organization WH. Update on the E-2020 initiative of 21 malaria-eliminating countries: report and country briefs. World Health Organization; 2018. WHO Document Code: 978-92-4-006351-6

7. Organization WH. Report of the first meeting of the WHO Diagnostic Technical Advisory Group for Neglected Tropical Diseases, Geneva, Switzerland, 30–31 October 2019: World Health Organization; 2020. https://books.google.com/books

8. Mengistu G, Diro E. Treatment outcome of severe malaria in adults with emphasis on neurological manifestations at Gondar University Hospital, north west Ethiopia. Ethiopian Journal of health development. 2006;20(2):106–11. 10.4314/ejhd.v20i2.10020

9. Gerbi A, Hassen M, Kebede W, Debella L, Bajaro M. Prevalence and pattern of severe malaria among adults in Jimma University Specialized Hospital, Jimma, Southwest Ethiopia: A three years retrospective study. Journal of Science and Sustainable Development. 2021;9(1):1–11. 10.20372/au.jssd.9.1.2021.0250

10. Ayele DG, Zewotir TT, Mwambi HG. Prevalence and risk factors of malaria in Ethiopia. Malaria journal. 2012;11:1–9. 10.1186/1475-2875-11-195

11. Kendie FA, Hailegebriel W/kiros T, Nibret Semegn E, Ferede MW. Prevalence of Malaria among Adults in Ethiopia: A Systematic Review and Meta-Analysis. Journal of tropical medicine. 2021;2021(1):8863002. 10.1155/2021/8863002

12. Mulugeta A, Assefa A, Eshetie A, Asmare B, Birhanie M, Gelaw Y. Six-year trend analysis of malaria prevalence at University of Gondar Specialized Referral Hospital, Northwest Ethiopia, from 2014 to 2019. Scientific Reports. 2022;12(1):1411. 10.1038/s41598-022-05530-2

13. Yimer M, Hailu T, Mulu W, Abera B, Ayalew W. A 5 year trend analysis of malaria prevalence with in the catchment areas of Felegehiwot referral Hospital, Bahir Dar city, northwest-Ethiopia: a retrospective study. BMC research notes. 2017;10:1–4. 10.1186/s13104-017-2560-6

14. Alemu A, Tsegaye W, Golassa L, Abebe G. Urban malaria and associated risk factors in Jimma town, south-west Ethiopia. Malaria journal. 2011;10:1–10. 10.1186/1475-2875-10-173

15. Gelaw B, Assefa S, Teklu T, Berhane W, Alem M, Sheferaw Y. Audit on the Laboratory Diagnosis of Malaria Parasites at the University of Gondar Hospital Laboratory, Northwest Ethiopia. Ethiopian Journal of Health and Biomedical Sciences. 2010;3(I):53–8. 10.20372/ejhbs.v3iI.172

16. Organization WH. Parasitological confirmation of malaria diagnosis: report of a WHO technical consultation, Geneva, 6-8 October 2009. 2010. WHO Publication Number: 978-92-4-159755-3

17. Lenicek Krleza J, Dorotic A, Grzunov A, Maradin M. Capillary blood sampling: national recommendations on behalf of the Croatian Society of Medical Biochemistry and Laboratory Medicine. Biochemia medica. 2015;25(3):335–58. 10.11613/BM.2015.034

18. Münster M. The role of the peripheral blood smear in the modern haematology laboratory. SEED haematology. Sysmex. February 2013. 2013. https://www.sysmex.co.jp/en/

19. Organization WH, UNICEF. Microscopy for the detection, identification and quantification of malaria parasites on stained thick and thin blood films in research settings (version 1.0): procedure: methods .manual. 2015. ISBN: 978 92 4 154921 9

20. Organization WH. Basic malaria microscopy: World Health Organization; 2010. ISBN: 978 92 4 154791 8(Part 2)

